# A Fresh View at Sports PSM-Systems

**DOI:** 10.1101/2023.01.05.23284213

**Authors:** Vladimir Savostyanov, Alexander Kobelev

## Abstract

**Background/Objectives:** The purpose of the proposed study is disclosure of correlations between the dynamics of the Breathe Rate (BR) and Heart Rate (HR) when performing the intermittent physical operation at maximum power on a cycle ergometer.

**Methods:** The stage on the study of the General functional athlete readiness (GFAR) performed using the sports standard “Red Engine” and the cycle ergometer in 16 volunteers (10 men, 6 women) whose average age was 21±1.17 years. To determine the sports potential of the volunteers in this study, we used own Coefficient of Anaerobic Capacity (CANAC Q, beats). Continuous recording of the heart rate and respiratory rate of volunteers in the maximum power sports test was carried out by the “RheoCardioMonitor” device with an athlete functional readiness module based on the method of Transthoracic electrical impedance rheography (TEIRG).

**Results:** The degree of correlation of functional indicators (M, HR_M_, GFAR) with CANAC Q for the group in full (n=80) occurred at a very high level, which confirmed the effectiveness of using the Coefficient of Anaerobic Capacity (CANAC Q) in assessing the general functional athlete readiness of the volunteers.

**Conclusions:** CANAC Q measured in “beats” of the heart and recorded very accurately using the method of transthoracic electrical impedance rheography (TEIRG). For this reason as promising sports PSM-system, CANAC Q can replace the methods for determining the functional athlete readiness by blood lactate concentration and maximum oxygen consumption.

## 1. Introduction

Physiological state monitoring (PSM-systems) is an IT systems used to record various physiological parameters of the human body.^1^ The purpose of the PSM-systems is to monitor the viability of a person in extreme conditions (diseases, injuries, extreme physical exertion, etc.)^2^

Generally accepted that the degree of the person’s efficiency in the terms of high intensity and tension (sports of high achievements, extreme activities, etc.) determined by complex interactions of the following components: 1) Health condition; 2) Physical (athletic) shape; 3) Technical (special) education; 4) Psychological motivation (morally volitional qualities).

For the clear understanding interactions of these components in athletic pedagogy, the term “Functional Athlete Readiness” is used.^3^

The concept of “Functional athlete readiness” (FAR) has a very complex and multifaceted context.^4^ The FAR may be defined as a relatively settled state of the organism, determined by the level of development of key functions required for the certain sport, as well as their specialized properties that directly or indirectly determine the effectiveness of the competitive activity.^5^

The FAR divided into two types:

a. General functional athlete readiness (GFAR), which characterizes the general physical development and endurance;
b. Special functional athlete readiness (SFAR) depending on the kinds of sport.

## 2. Methods

During 2015-2017 Bauman Moscow State Technical University together The University of Alabama (USA) was carried out a scientific project “A study of applicability of a breathing sensor PACT2.0 in determination of aerobic-anaerobic potential of professional athletes (junior ice hockey player)”. This study has been approved by the Institutional Review Board for the protection of human subject request for approval of research involving human subjects (IRB Protocol #15-024-ME 2015/12/14), as well as by the Decisions of the Ethics Committee of Bauman Moscow State Technical University No.1 from 2015/11/11 and No.2 from 2016/10/01).

The purpose of this study was disclosure of correlations between the dynamics of the Breath Rate (BR) and Heart Rate (HR) when performing the intermittent operation at maximum power on a cycle ergometer.

The study allowed to determine anaerobic capacity of sportsmen in the framework of their general functional athlete readiness and to evaluate feasibility of developing a sports methodology for diagnostics the state of syndrome chronical overexertion of the heart.

The necessity for this research determined by current requirements of the strategical item of a coach the information about how professional athletes in team kinds of sport (hockey, football, handball, basketball, etc.) match each other in speed-strength qualities with a determination of a possibility for their mutual substitution without loss of a game quality during competitive cycles.

Developed during this study the original PSM-system named the sports standard “Red Engine”. The “Red Engine” solves the global problem in the determination of the individual dynamic of changes in the levels of functional readiness of each sports team member, that allows effectively to maintain high levels of team efficiency in the whole during a competitive macrocycle.

The convenience of this sports functional testing is that it held directly in the conditions of a stadium without involving any complex diagnostic medical equipment.

The heart rate (HR) most often used as a load intensity evaluation criterion in sports. There is a linear relationship between heart rate and training intensity.^6^

Often, endurance trainings (aerobic exercises) performed by the athletes at the heart rate of about 180 beats per minute (bpm). However, for many athletes this heart rate significantly exceeds their personal aerobic-anaerobic transition area.

Therefore, for calculate training intensity and monitor an athlete’s functional state must use him Resting heart rate (HR), Maximum heart rate (HR_max_), Reserve heart rate (HR_R_) and Target heart rate of the transition to completely anaerobic zone (HR_M_).

HR_max_ is the maximum number of contractions that the heart may be made within one minute. After twenty years, HR_max_ gradually begins to decline by about one beat per year. So HR_max_ is calculated by the following formula^7^:

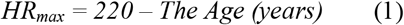

For the calculation of the load intensity is also used the method of heart rate reserve (HR_R_), which was developed by Finnish scientists Karvonen. HR_R_ is the difference between HR_max_ and HR rest (HR_0_):

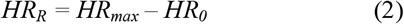

Knowing the HRR, you can calculate the target of the heart rate (HR_M_). Target heart rate (HR_M_) is the optimal heart rate, characterizing the transition in the clear anaerobic zone during the intensity of the performed exercise M (%):

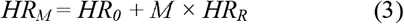

At the same time, knowing HR_0_ and HR_max_, according to the Karvonen’s formula, it is possible to calculate with what intensity (M) the athlete performs the exercise:

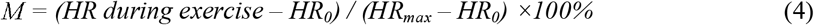

Thus,

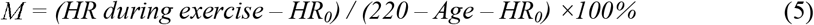

The sports standard “Red Engine” was created for a functional testing of the professional athletes during training camps and competitions. The basis of this functional testing is registration of different biometric and anthropometric indicators with the further complex of very difficult mathematical processing.

Based on results of the sports standard “Red Engine”, make an individual biomedical program and including recommendations (**Fig. 1**):

**Fig. 1.**
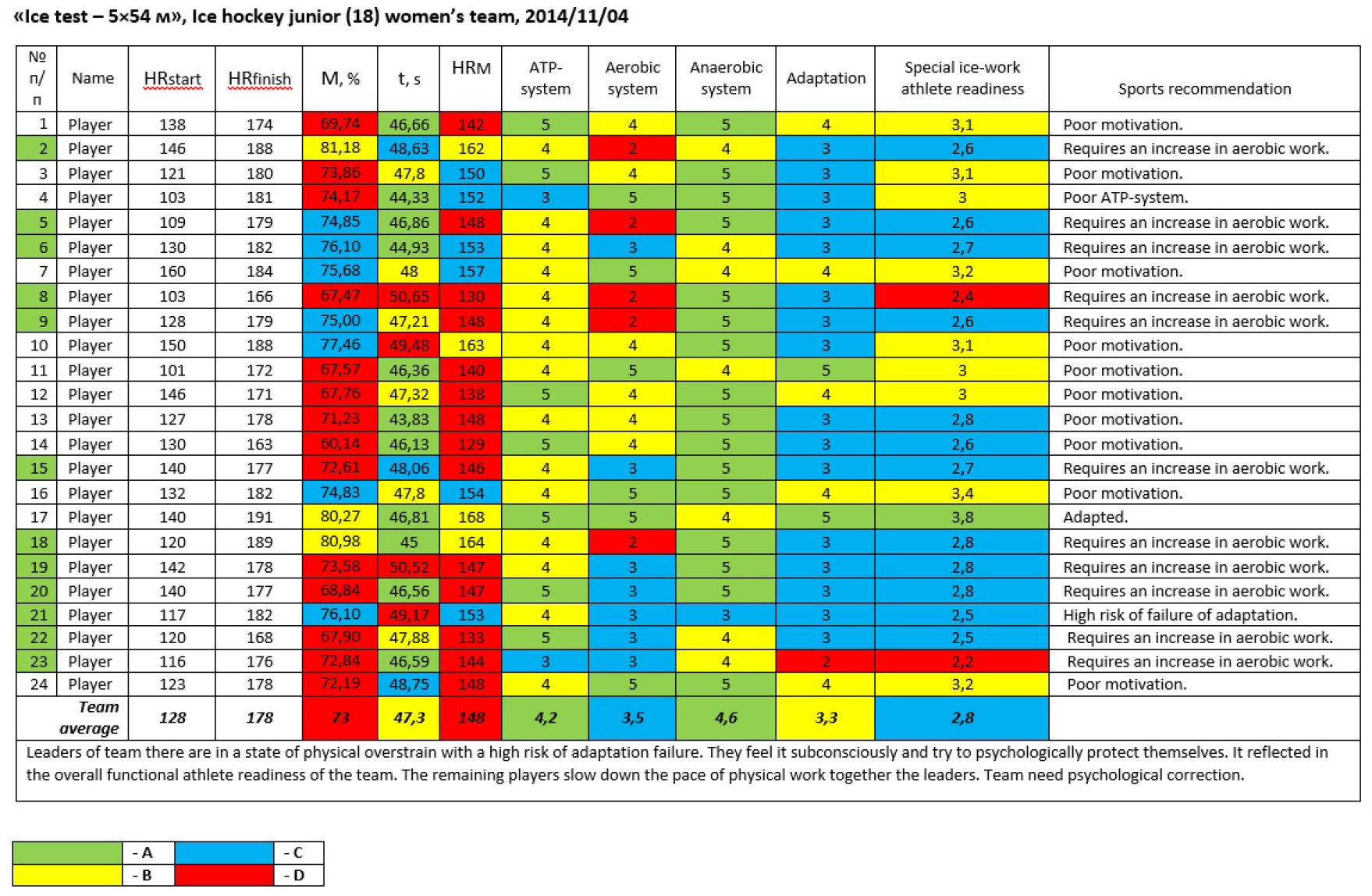
Pedagogical interpretation of the sports standard “Red Engine” results.

1. Additional work or rest during educational training camp;
2. Preventive measures;
3. Using of the selective methods of restoring the functions of ATP-system, Aerobic- and Anaerobic-system;
4. Plan of correction of a medical and biological providing and a pharmacological protection in extreme conditions of sport.

## 3. Results

The created sports standard “Red Engine” turned out to be very effective in the training of professional athletes. The “Red Engine” was validation by the methods for determining the athlete functional readiness by blood lactate concentration and maximum oxygen consumption.

However, it requires the use of special software that is not available to a wide audience of users. For this reason, we decided to simplify this method in the diagnostic variant of study only General functional athlete readiness.

The stage on the study of the General functional athlete readiness (GFAR) performed using the sports standard “Red Engine” and the cycle ergometer in 16 volunteers (10 men, 6 women) whose average age was 21±1.17 years. All volunteers were members of our University sports club.

Using test based on the achievement by athletes of the maximum power of muscle load in five series of 45″, in which the heart rate rises to maximum values, followed by recovery pauses of 90″.

To determine the sports potential of the volunteers in this study, we used own Coefficient Anaerobic Capacity (CANAC Q, beats):

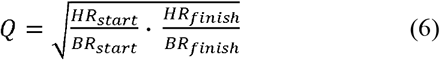

Continuous recording of the heart rate and respiratory rate of volunteers in the maximum power sports test was carried out by the “RheoCardioMonitor” device with an athlete functional readiness module based on the method of Transthoracic electrical impedance rheography (TEIRG).^8–12^

The method of Transthoracic electrical impedance rheography, which allows registering (allows to register) the frequency characteristics of the volunteer’s bioimpedance response on the sport test of maximum power, thereby simultaneously recording the characteristics of his productive work of the heart channel, the effectiveness of the blood gas transport function, as well as to reliably assess its reactivity and adaptive potential.^13^

In technical terms, the “ReoCardioMonitor” system represents a two-channel impedance-measuring converter, a patient’s cable system. The first channel designed to measure the transthoracic impedance of the chest (base and pulse components, impedance-breathing pattern) with an ECG measurement channel from the same impedance electrodes. The transthoracic channel used to calculate stroke output and cardiac output. The second channel measures breathing pattern.^14^

The measurement method is tetrapolar. A source that creates an alternating current of high frequency connected to the current electrodes. The signal recorded from potential electrodes. With an increase in the distance between the current and potential electrodes, their influence on each other decreases and the accuracy of the measurement result increases.^13^

The use of such a scheme for applying electrodes ensures greater uniformity of the current passing through the investigated area and reduces the effect on the measurement result of resistance variations at the electrode-skin interface.^14^

Contact with the patient’s body was provided with disposable ECG electrodes of the White Sensor 4500 made “Ambu” company, electrode size 50×48 mm. The inner electrodes, marked in blue in the diagram, are measuring and registering the voltage change ΔU(t), which is used to calculate and register the impedance signal dZ. External electrodes are required to pass AC ∼ I with high frequency and low amplitude. The change in impedance is recorded as a function of time Z(t).

Classification of the studied functional indicators shown in **Tab. 1**.

**Table 1.** Classification of the studied functional indicators.

| Indicators | Result | Score |
| --- | --- | --- |
| The intensity of the performed exercise (M, %)* | $\geq 85$ | A |
|  | 80-84.9 | B |
|  | 75-79.9 | C |
| | $\leq 74.9$ | D |
| Target heart rate of the transition to completely anaerobic zone (HR<sub>M</sub>, bpm)* | $\geq 171$ | A |
|  | 161-170 | B |
|  | 151-160 | C |
| | $\leq 150$ | D |
| General functional athlete readiness (GFAR, units)* | $\geq 3.5$ | A |
|  | 3.0-3.4 | B |
|  | 2.5-2.9 | C |
| | $\leq 2.4$ | D |
| Coefficient Anaerobic Capacity (CANAC Q, beats) | $\geq 10.1$ | A |
|  | 7.5-10 | B |
|  | 5.5-7.4 | C |
| | $\leq 5.4$ | D |

\* - classification of Russian Ice Hockey Federation.<sup>4</sup>

## 4. Discussion

The results of the study (**Tab. 2**) measure the low levels of the General Functional Athlete Readiness of volunteers: on average GFAR=2.5 units (Score “C”) were in all test series. Professional athletes must have a GFAR of at least 3.5 units.

**Table 2.** Results of the study of the General Functional Athlete Readiness with using the method “Red Engine” in all series.

| Indicators | 1st series | 2nd series | 3rd series | 4th series | 5th series |
| --- | --- | --- | --- | --- | --- |
| HR<sub>start</sub>, bpm | 102,8±13,6 | 137,1±15,7 | 144,4±17,9 | 148,2±15,1 | 152,1±16,2 |
| HR<sub>finish</sub>, bpm | 158,4±15,7 | 167,9±14,7 | 173,1±11,9 | 173,9±11,6 | 176,8±11,9 |
| BR<sub>start</sub>, rpm | 17,1±6,9 | 19,4±4,7 | 22,1±5,7 | 23,4±4,7 | 24,1±6,0 |
| BR<sub>finish</sub>, rpm | 33,7±6,5 | 35,1±5,1 | 37,6±7,2 | 39,5±7,9 | 40,0±7,6 |
| M, % | 66,7±8,4 | 65,9±7,2 | 67,7±5,9 | 67,2±7,1 | 68,4±7,0 |
| HR<sub>M</sub>, bpm | 129,5±18,1 | 134,7±17,4 | 134,7±17,4 | 140,1±15,7 | 143,4±15,7 |
| GFAR, units | 2,51±0,70 | 2,48±0,63 | 2,59±0,64 | 2,54±0,60 | 2,61±0,57 |
| CANAC Q, beats | 5,7±1,29 | 5,9±0,87 | 5,7±1,13 | 5,4±0,92 | 5,5±0,99 |

The value of the individual intensity (power) of the exercise (M) did not reach 70% (Score “D”) in any of the test series. The threshold value of anaerobic metabolism (HR_M_) does not go beyond 145 bpm (Score “D”). The professional sports quality level must at least 171 bpm.

Positive correlations of functional indicators (M, HR_M_, GFAR) with the Coefficient of Anaerobic Capacity (CANAC Q) were registered in the first two test series. However, during continuing series of work with increased power the correlations between the functional indicators began to destroy due to the pronounced fatigue of the volunteers (**Tab. 3**).

**Table 3.** The degree of correlation of functional indicators with CANAC Q in all series of the study.

| Indicators | 1st series | 2nd series | 3rd series | 4th series | 5th series |
| --- | --- | --- | --- | --- | --- |
| M, % | r=0,50 | r=0,41 | r=0,18 | r=0,09 | r=0,02 |
|  | p<0,01 | p<0,01 | p>0,05 | p>0,05 | p>0,05 |
| HR<sub>M</sub>, bpm | r=0,55 | r=0,40 | r=0,19 | r=0,08 | r=0,04 |
|  | p<0,01 | p<0,01 | p>0,05 | p>0,05 | p>0,05 |
| GFAR, units | r=0,53 | r=0,51 | r=0,38 | r=0,29 | r=0,33 |
|  | p<0,01 | p<0,01 | p<0,05 | p>0,05 | p>0,05 |

The average maximum heart rate (HR_finish_) of volunteers in all series could not overcome the mark of 170.0±14.8 bpm, although according to the Karvoner’s formula, their maximum heart rate must reach 199 bpm. The heart rate start (HR_start_) was 136.9± 23.7 bpm.

The maximum breath rate (BR_finish_) also turned out to be excessively high 37.1±7.4 rpm. At the same time the breath rate start (BR_start_) was 21.2 ± 6.2 rpm, which also indicates a very weak physical working capacity of this group of volunteers.

The average CANAC Q of volunteers in all series could not overcome the mark of 5.6±1.07 beats (Score “C”).

The degree of correlation of functional indicators (M, HR_M_, GFAR) with CANAC Q for the group in full (n=80) occurred at a very high level (**Tab. 4**), which confirmed the effectiveness of using the Coefficient Anaerobic Capacity (CANAC Q) in assessing the General functional readiness of an athlete.

**Table 4.** The degree of correlation of functional indicators with CANAC Q for group in full.

| Indicators | Group in full (n=80) | Degree of correlation<br>with CANAC Q |
| --- | --- | --- |
| M, % | $67,2 \pm 7,2$ | $r=0,241$; $p<0,01$ |
| $HR_M$, bpm | $137,5 \pm 17,1$ | $r=0,237$; $p<0,01$ |
| GFAR, units | $2,55 \pm 0,63$ | $r=0,406$; $p<0,01$ |

## 5. Conclusion

Thus, the results of the study showed a very low average level of the General Functional Athlete Readiness among university sports club volunteers when using the sports standard “Red Engine” (GFAR=2.55±0.63 units with the required level of more than 3.7 units). Moreover, the same result was obtained using a new sports indicator – the Coefficient Anaerobic Capacity (CANAC Q), which amounted to 5.6 ± 1.07 beats with the required level of more than 10.1 beats.

The fundamental idea of CANAC Q is calculating of the ratio of increase in heart rate (HR) to increase of breath rate (BR) during intensive physical activity: than less BR spent to achieve maximum heart rate, thereby will be higher the anaerobic capacity of athletes. CANAC Q measured in “beats” of the heart and recorded very accurately using the method of transthoracic electrical impedance rheography (TEIRG). For this reason, as promising sports PSM-system, CANAC Q can replace the methods for determining the athlete functional readiness by blood lactate concentration and maximum oxygen consumption.

## Data Availability

All data produced in the present study are available upon reasonable request to the authors

http://www.healthscreens.guru/

## Author statement

The authors agree with the content of the manuscript and approve of its submission to the Journal of Exercise Science & Fitness (JESF). We confirm that the manuscript has not been previously published (except in abstract form) and is not being concurrently submitted elsewhere, and will not be submitted to another journal before a final editorial decision from JESF is rendered.

## Acknowledgements

We express our sincere gratitude to all the participants (volunteers) of our experiment. Without them, this study would not have been possible.

